# A systematic review of methylome-wide associations with anxiety disorders

**DOI:** 10.1101/2025.07.13.25331464

**Authors:** Sarah J. Ingram, Lea Zillich, Miriam A. Schiele, Psychiatric Genomics Consortium Anxiety Epigenetics Workgroup, Katharina Domschke, John M. Hettema, Shaunna L. Clark

## Abstract

Anxiety disorders (ANX) are a prevalent public health burden that significantly impair daily functioning and decrease quality of life. A growing body of research suggests that DNA methylation (DNAm), an epigenetic modification that can impact gene expression, may be altered in ANX. The current review used a systematic approach to identify and synthesize the literature regarding methylome-wide association studies (MWASs) of ANX in humans. We screened 804 articles returned by a search in PubMed in May 2025 and identified 12 studies for inclusion. All included studies examined ANX-associated DNAm in blood. In total, 2,023 DNAm sites corresponding to 985 genes were significantly associated with ANX. No DNAm sites significantly replicated across studies and four nominally replicated. This is likely a result of a lack of replication attempts, small sample sizes, and differences in data analysis choices. Findings suggest that ANX-associated DNAm may promote dysregulation of immune and inflammatory processes, some possibly sex-dependent. Collectively, the findings from studies included in this review provide preliminary evidence of ANX-related alterations to DNAm in whole blood and multiple blood cell-types. Future MWASs of ANX could benefit from larger sample sizes, a standardized analytic pipeline, longitudinal study designs, and the examination of DNAm in additional cell-types and tissues.

## Introduction

Anxiety is a normative response to perceived and actual threats that, when dysregulated, can transform into a debilitating disorder characterized by worry and fear so excessive it is disturbing to one’s daily life (Craske et al., 2009). Anxiety disorders (ANX) are prevalent with over 300 million people worldwide experiencing one or more ANX in 2019 (Javaid et al., 2023). The Diagnostic and Statistical Manual of Mental Disorders 5^th^ edition (DSM-5; (American Psychiatric Association, 2013)) defines seven primary ANX with the prevalence of each ranging as follows: generalized anxiety disorder (GAD) (3%), panic disorder (2-3%), social anxiety disorder (SAD) (7%), specific phobia (8-12%), agoraphobia (1-2%), separation anxiety (2%), and selective mutism (<1%) (American Psychiatric Association, 2013). Research into the pathoetiology of ANX lags somewhat behind other mental disorders despite its high prevalence and widespread impacts. Heterogeneous clinical presentations and extensive comorbidity with other mental disorders complicate investigations of the etiology of ANX (Doering et al., 2019; Drzewiecki & Fox, 2024; Kessler et al., 2005). Like most medical phenotypes, risk for ANX is attributable to a combination of genetic and environmental sources (Hettema et al., 2005; Levey et al., 2020). ANX are highly polygenic with an estimated 20 to 60% of the variance attributed to common genetic factors (Gottschalk & Domschke, 2017; Hettema et al., 2005; Purves et al., 2020). Environmental factors, such as adverse life events, also contribute and may interact with genetic factors to confer risk for ANX (Danielsdottir et al., 2024; Nelson et al., 2002). However, biological mechanisms through which these risk factors and their interplay exert their effects remain elusive.

Suggested biological mechanisms include epigenetic modifications that influence gene expression without altering the underlying DNA nucleotide sequence. We focus on DNA methylation (DNAm), the most studied epigenetic modification in humans. DNAm involves the addition of a methyl group to the carbon 5 position of a cytosine base and is most commonly found in the sequence context CpG (i.e., cytosine-phosphate-guanine). DNAm is largely conserved throughout the human population and maintainable across cell division, making DNAm integral to proper cell and tissue functioning (Ehrlich & Lacey, 2013; Farlik et al., 2016; Houseman et al., 2015; Loyfer et al., 2023). DNAm alterations occur in response to genetic and environmental triggers which change overtime across cells and tissues, suggesting DNAm may play a role in dynamically adapting to changing environments (Wiegand et al., 2021). It is thought DNAm may confer increased risk for ANX by causing a maladaptive response to genetic and environmental triggers (Schiele et al., 2020). A growing body of research has found evidence of altered DNAm in depression and post-traumatic stress disorder which are closely related to ANX (Li et al., 2019; Morrison et al., 2019); however DNAm alterations in ANX are far less studied and understood.

Early DNAm studies of ANX focused on CpGs in promoter regions of candidate genes involved in stress response and neurotransmission and reported significant associations with ANX (Chagnon et al., 2015; Domschke et al., 2012; Womersley et al., 2022; Ziegler et al., 2015). Candidate gene DNAm studies have been broadly criticized for their lack of replicability, which is likely due to false positive associations (Shabalin et al., 2015). Several reasons have been suggested for this including small sample sizes which lack power to detect effects, no correction for multiple testing, and lack of adjustment for relevant confounders. For example, candidate gene DNAm studies typically do not control for differences in cell-type composition despite known impacts on comparisons of DNAm between samples (Houseman et al., 2015; Qi & Teschendorff, 2022). Not controlling for cell-type composition produces inflated test statistics and, therefore, a large number of seemingly significant associations. Thus, significant findings in candidate gene DNAm studies may reflect ANX-related changes in cell-type composition rather than true associations (Houseman et al., 2015). Further, candidate gene approaches typically examine a small number of CpGs in the promoter region of one gene with a putative *a priori* relationship to ANX. This narrow focus disregards CpGs located outside of promoter regions and has limited discovery of ANX-associated sites to the handful of genes examined. Such limited approaches put DNAm studies in danger of repeating the long, largely unsuccessful history of psychiatric candidate gene association studies (Border et al., 2019; Howe et al., 2016; Johnson et al., 2017; Sullivan, 2007).

Methylome-wide association studies (MWAS) analyzing associations between observable traits and CpGs across the methylome are preferred, as they can overcome some limitations of candidate gene studies. MWAS are hypothesis-free and not limited to promoter regions of specific genes, allowing for a more comprehensive investigation of the methylomic landscape in ANX. Routine inclusion of relevant covariates, such as cell-type composition, and correction for multiple testing further decreases the likelihood MWAS findings are false positives (Campagna et al., 2021). These advances mirror those seen over the past decade in psychiatric genetic association studies (Levey et al., 2020; Purves et al., 2020). Reviews of MWASs of depression and PTSD have been conducted (Li et al., 2019; Morrison et al., 2019), yet an in-depth examination of the methylomic landscape in ANX has not been investigated. Here, we conduct a systematic literature review to identify MWAS of ANX, describe study characteristics, and synthesize major themes among findings.

## Methods

### Search Strategies and Study Selection

This systematic review was conducted in accordance with the Preferred Reporting Items for Systematic Reviews and Meta-Analyses (PRISMA) guidelines (Page et al., 2021). ANX was defined as a diagnosis or symptom count of any of the seven primary ANX. We included studies assessing DNAm associations with additional phenotypes, but only reviewed portions relevant to the scope of this review. To be included, studies must have evaluated DNAm on a methylome-wide scale in humans, used a phenotype of ANX as defined above, and published in English in a peer-reviewed journal (i.e., preprints were not considered). We also only considered studies correcting for cell-type composition to ensure findings reviewed reflect true associations instead of ANX-related differences in cell-type composition.

Studies were identified by searching PubMed for empirical articles meeting inclusion criteria in May 2025. The search term used was (“anxiety” OR “generalized anxiety” OR “specific phobia” OR “social phobia” OR “social anxiety” OR “agoraphobia” OR “panic” OR “panic disorder” OR “separation anxiety” OR “mutism”) AND (“methylome” OR “methylome wide” OR “methylome-wide” OR “epigenome” OR “epigenome wide” OR “epigenome-wide” OR “methylation” OR “DNA methylation”). Titles and abstracts were reviewed by SJI and SLC. Discrepancies were resolved by consensus. Duplicates, reviews, pre-prints, retractions, studies conducted in non-human or non-biological models, studies not examining ANX, and studies not examining DNAm on a methylome-wide scale were excluded (Figure 1).

**Figure 1.**
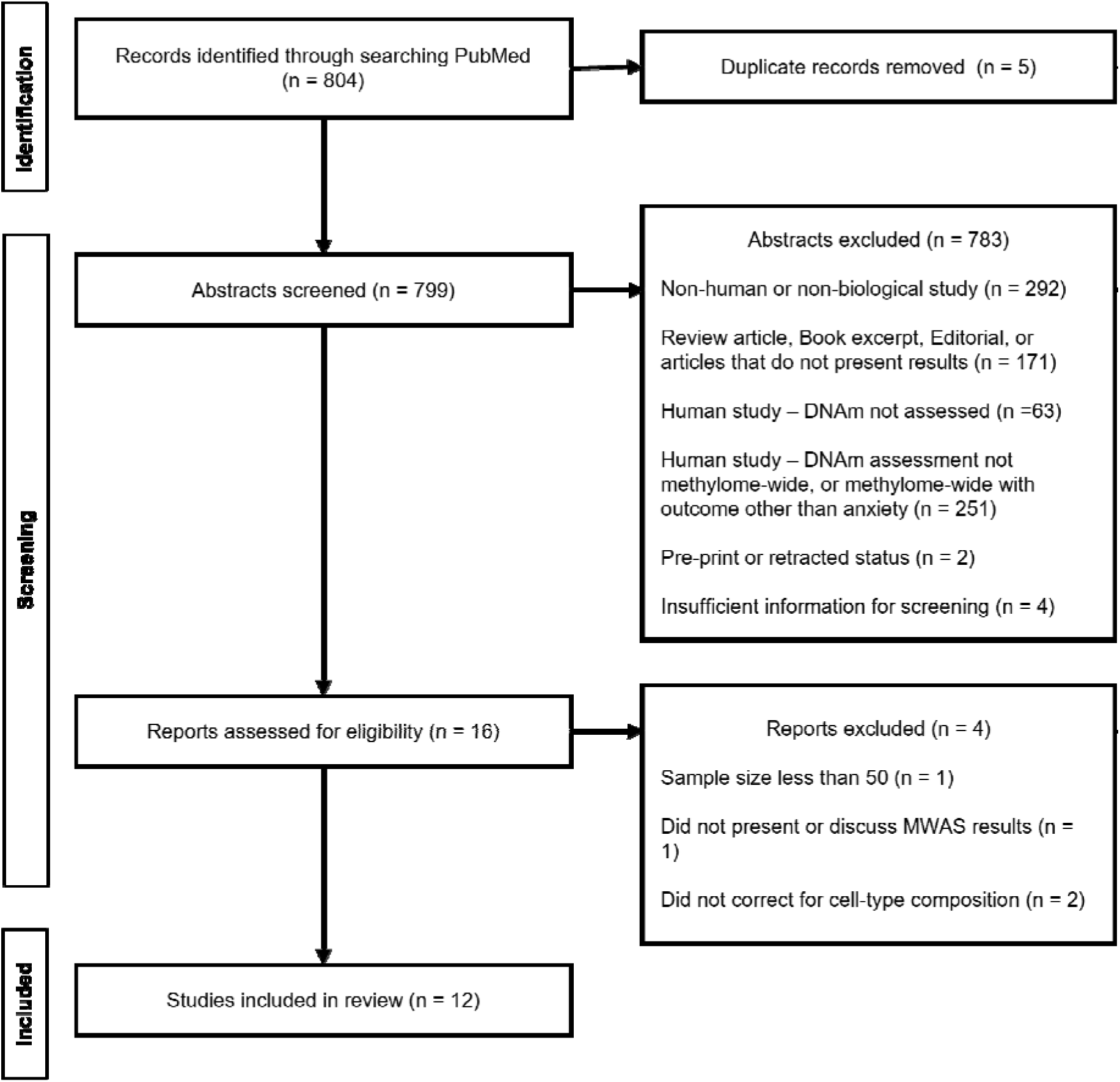
PRISMA flow diagram.

For the sixteen studies passing initial screening, data were extracted on study sample characteristics, study design, covariates included, multiple testing correction methods, main MWAS findings including the location of significant findings, and findings from replication attempts in independent samples. Four studies were excluded for the following reasons: 1) conducted a MWAS in six participants (Alisch et al., 2017), which is drastically underpowered to detect effects in a MWAS (Mansell et al., 2019), 2) only evaluated downstream impacts of ANX-associated DNAm on biological pathways and did not report main MWAS findings (Bortoluzzi et al., 2018), and 3) did not account for cell-type composition (Zhou et al., 2015; Zou et al., 2023).

We conducted a cross-study replication to assess the reproducibility of ANX MWAS findings and identify potential CpGs or genes warranting further investigation. For this purpose, we extracted information on CpGs from each study that were nominally significant (p < 0.05). A CpG was considered significantly replicated if it met methylome-wide significance criteria in more than one study and had the same direction of effect. Sites were considered nominally replicated if they met methylome-wide significance in one study and nominal significance defined as p <0.05 in another, also with the same direction of effect. We also assessed if there was replication on the gene-level by examining if CpGs that met methylome-wide significance criteria in more than one study were annotated to the same gene. Annotations provided by the original studies were used for this purpose.

## Results

The search yielded 804 articles of which 12 met criteria for inclusion (Ciuculete et al., 2018; Domschke et al., 2025; Emeny et al., 2018; Guo et al., 2022; Hettema et al., 2023; Iurato et al., 2017; Kwon et al., 2025; Ohi et al., 2024; Petersen et al., 2020; Shimada-Sugimoto et al., 2017; Wiegand et al., 2021; Ziegler et al., 2019). These studies are summarized in Table 1. All studies evaluated ANX-associated DNAm cross-sectionally with 11 studies having a case-control design. Discovery samples ranged in size from 96 to 1,522 participants. Most studies examined a single ANX or its symptoms, namely panic disorder, GAD, and SAD. The remaining studies evaluated associations with composite phenotypes of multiple ANX, either current or lifetime, though the ANX included in each study varied. The majority of participants across studies were of European ancestry, however multiple Asian sub-populations were also represented. Participants from all studies were mostly female consistent with the sex imbalance in anxiety prevalence. Adolescents were considered in one study while adults were the focus of the remaining eleven.

**Table 1:**
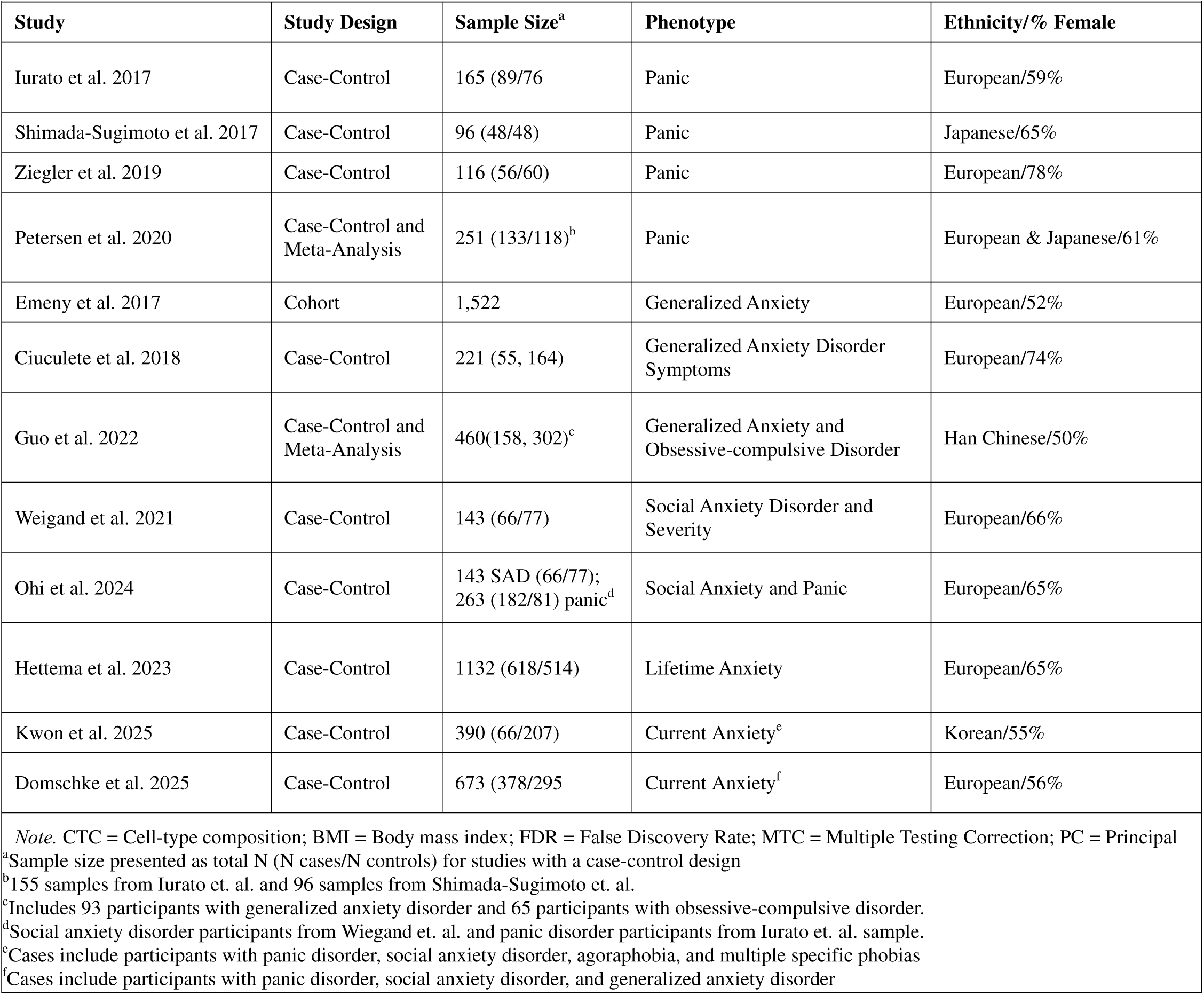
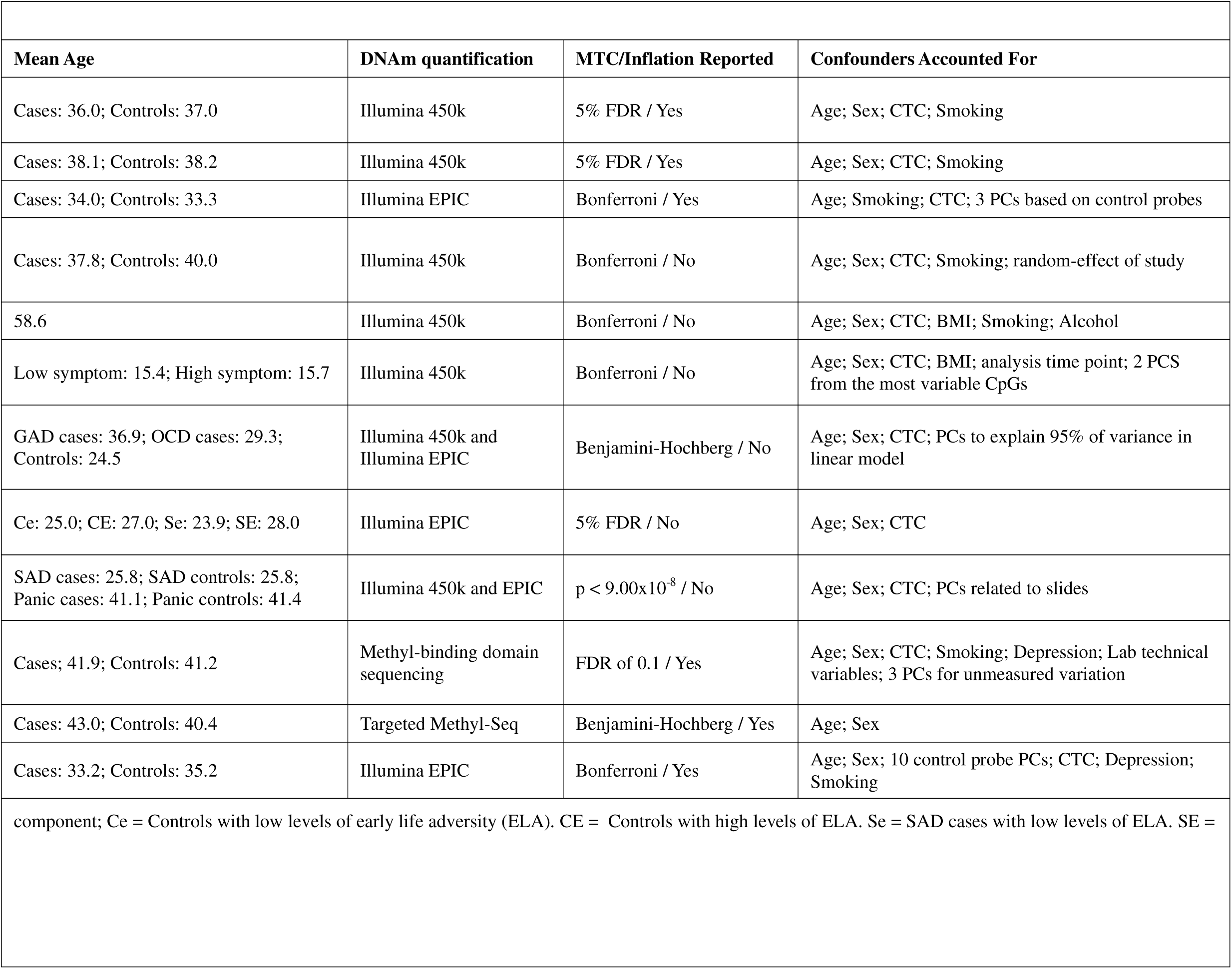
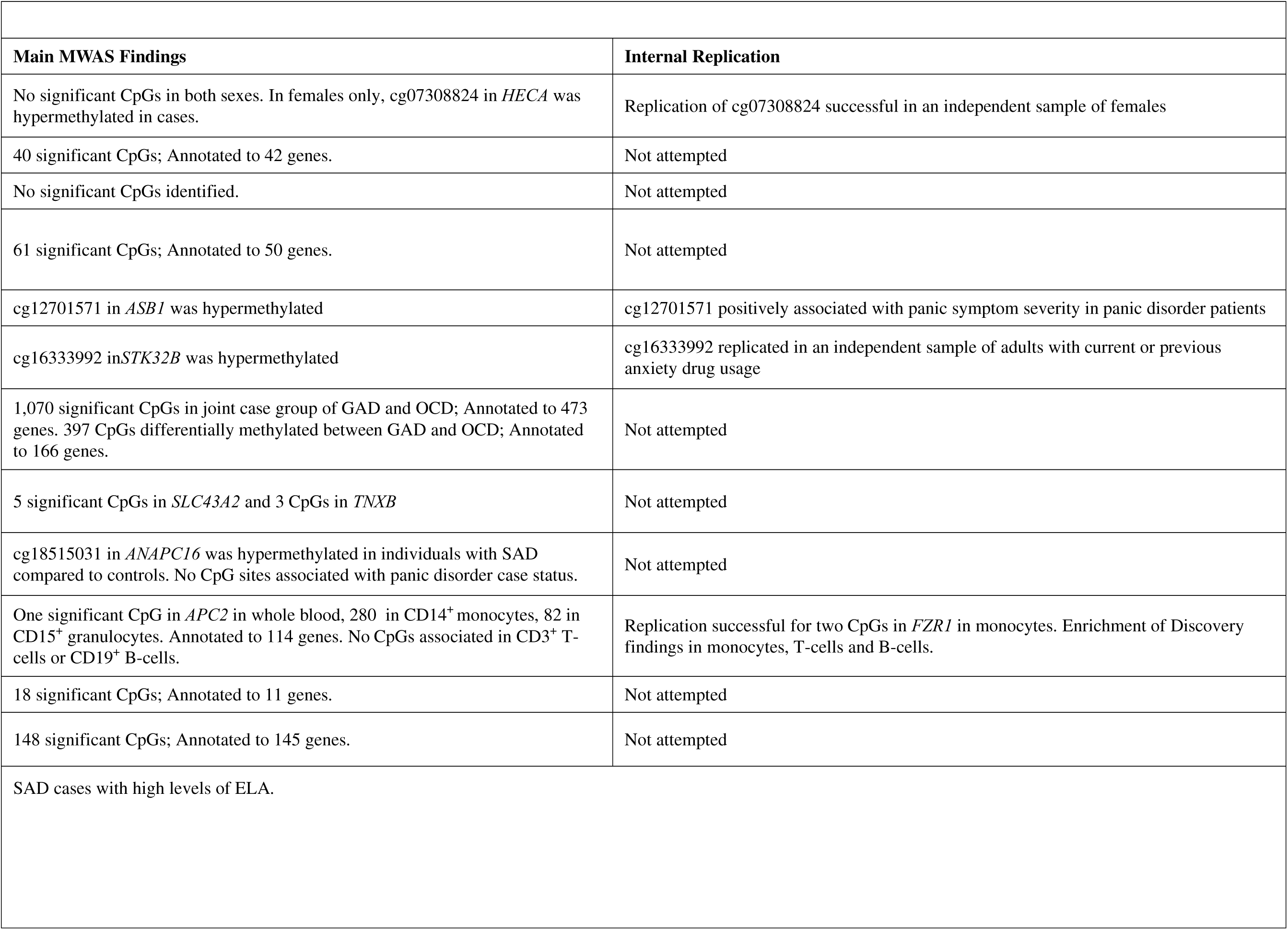
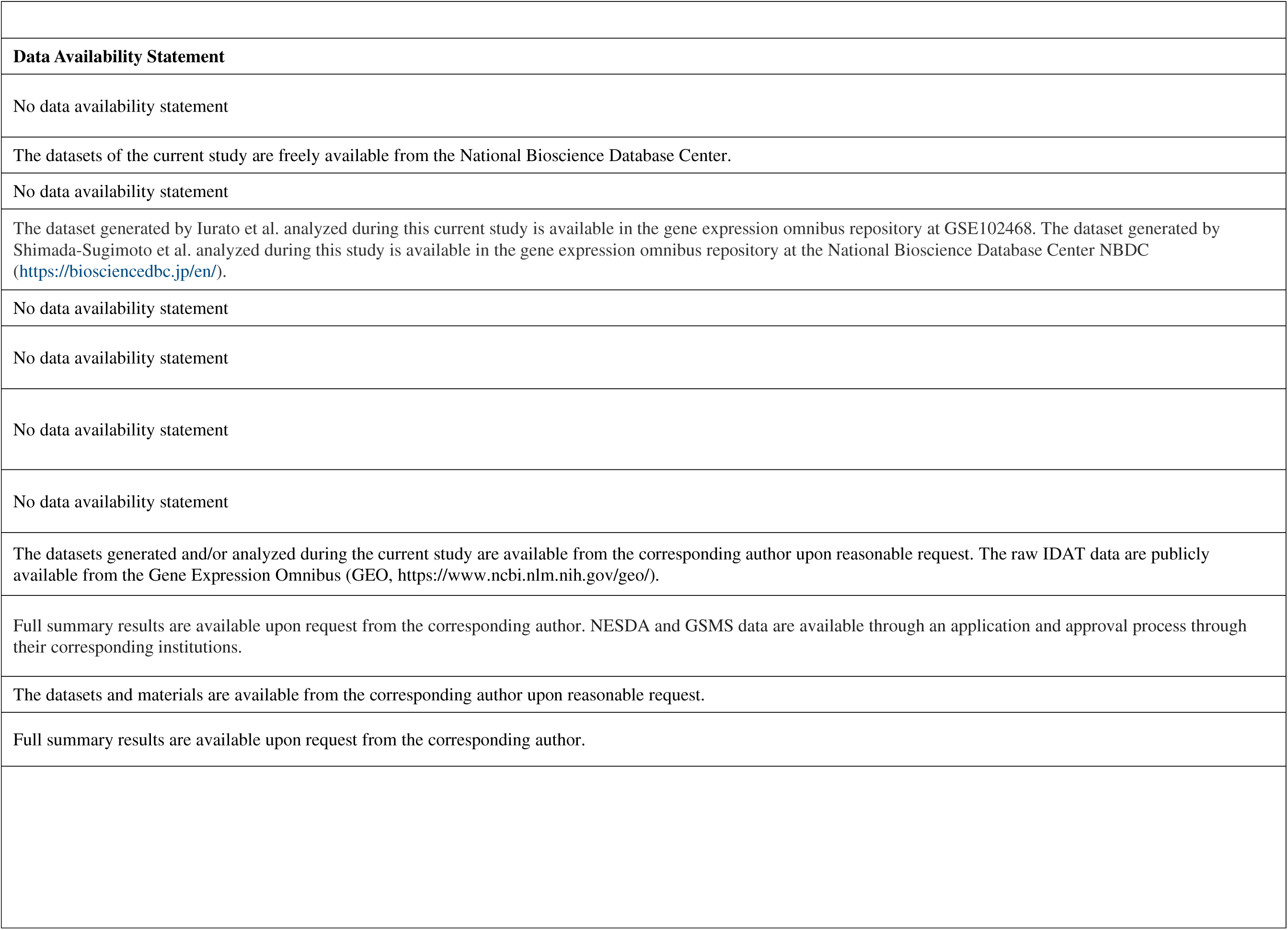
Methylome-wide association studies of ANX meeting inclusion criteria.

MWAS findings reflect ANX-associated DNAm in blood only, as no studies used alternate tissues, and only one study conducted additional evaluations in different blood cell-types. DNAm was quantified with array-based technologies in 10 studies and sequencing-based methods in the remaining two. Multiple testing correction was performed in all studies, though the choice of correction method and threshold for declaring methylome-wide significance varied. Although the covariates included was study-specific, potential confounding from age, sex, and cell-type composition was consistently controlled for across all studies. Smoking was accounted for in seven studies. Four studies attempted to replicate their significant findings in an independent sample, with four CpGs replicating within-study. Potential test statistic inflation was evaluated in five studies and six studies provided statements regarding the availability of raw data and/or summary statistics.

In total, 2,023 CpGs reached methylome-wide significance with ANX after correction for multiple testing, which corresponded to 985 unique genes (Table S1). The literature collectively suggests ANX are associated with altered DNAm; however cross-study replicability was minimal as no CpGs significantly replicated. However, cg03019505, cg25372841, and cg25947600 met methylome-wide significance thresholds in more than one study but had opposite directions of effect (Domschke et al., 2025; Guo et al., 2022) (Table 2). Four CpGs nominally replicated. Additional CpGs may meet nominal replication criteria, however we could not evaluate this possibility as some studies only reported methylome-wide significant findings or findings related to candidate genes. A full list of all CpGs reported by the studies reviewed here is in Table S2. At the gene-level, 20 genes annotated to a significant finding in more than one study (Table S3).

**Table 2:**
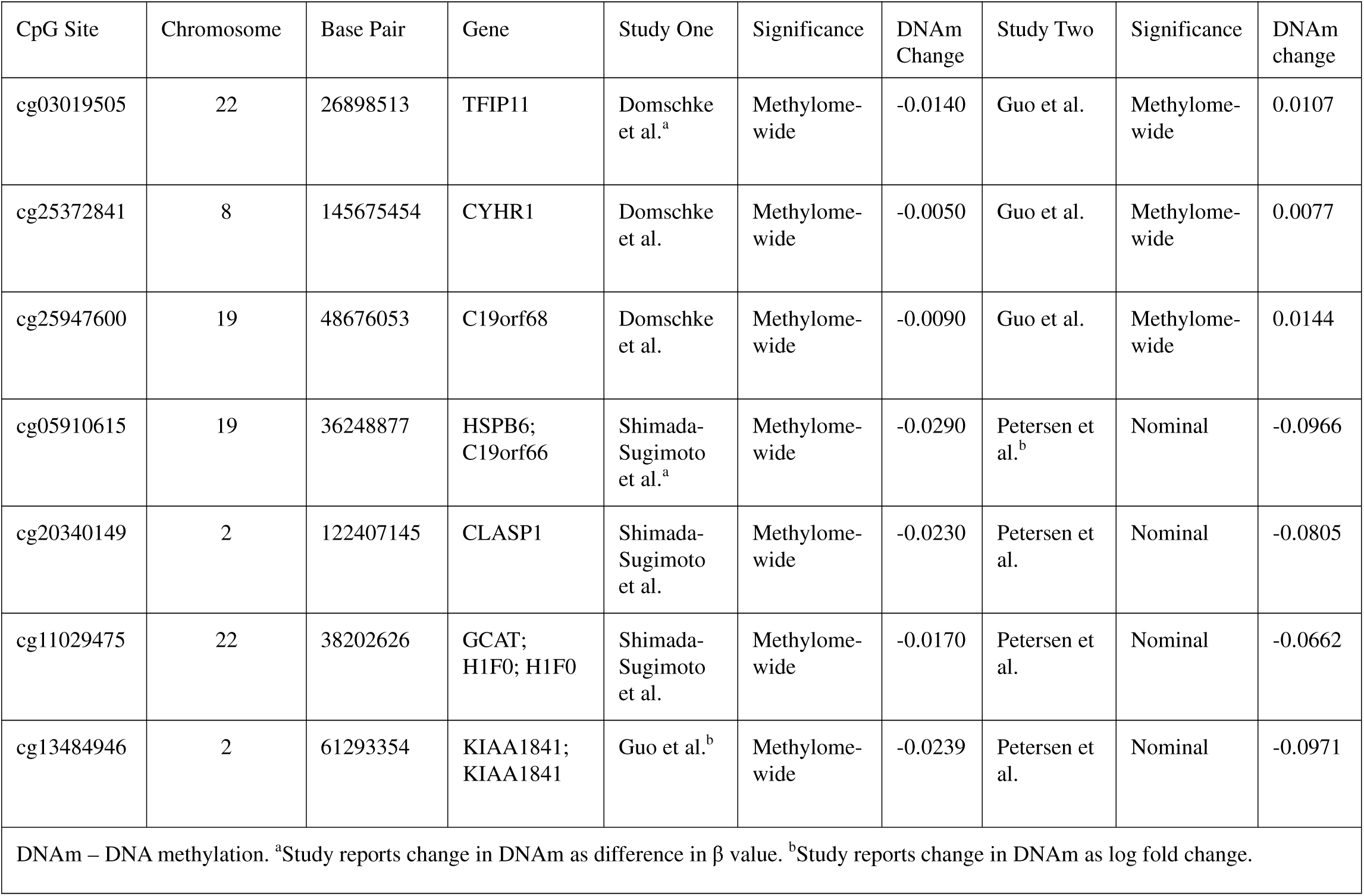
Partial Replication of CpG sites across studies.

## Discussion

This systematic review examined methylome-wide ANX-associated DNAm, focusing on 12 empirical studies in humans. Notable strengths of the literature included consistent use of multiple testing correction and inclusion of relevant confounders (e.g., age, sex, cell-type composition). Anxiety phenotypes were clearly defined in all studies, which is critical for drawing meaningful conclusions. Findings from reviewed studies provide preliminary evidence of ANX-associated DNAm, yet we identified an overall lack of replicating associations across studies. Three CpGs reached methylome-wide significance in more than one study, but their associations were in opposite directions, making it difficult to clearly identify specific CpGs warranting further investigation.

Low cross-study replicability could be driven by differences in DNAm platform coverage, a lack of replication attempts, and small discovery sample sizes. DNAm arrays and sequencing methods vary in coverage and quality of measurement, which may have hindered replication attempts. Additionally, only a handful of studies attempted to replicate their discovery findings in independent samples, making it difficult to discern true findings from potential false positives. Most studies used samples of less than 250 participants that were likely underpowered to detect the smaller DNAm effects expected in anxiety (Mansell et al., 2019). Sample sizes could be increased by utilizing large-scale biobanks or combining datasets using meta-analysis. By meta analyzing two panic disorder MWASs, Petersen et al. (2020) uncovered associations not detected in the individual MWASs. For the purposes of this review, a meta-analysis was not possible as the availability of raw data or summary statistics was inconsistent. Additionally, the heterogeneity in quality control, pre-processing, and analytic choices made conducting a meta-analysis of included studies inappropriate according to PRISMA guidelines. A more standardized MWAS analytic pipeline would homogenize data across studies and eliminate variation introduced by study-specific analysis choices. There are no agreed upon ‘best’ methods for analyzing MWASs data, however recommendations for best practice in MWAS study design are presented elsewhere (Campagna et al., 2021).

Study-specific analysis choices may have also affected replicability. Consistent correction for multiple testing across studies effectively minimized false positives, but multiple methods were used. Some true effects may have gone undetected in studies employing more conservative thresholds. Inconsistent inclusion of relevant confounders likely impacted replicability further. Depression is highly comorbid and shares many underlying risk factors with anxiety (Kalin, 2020), and as such some DNAm patterns may also be shared. Yet only 2 studies controlled for the potential influence of depression on their analyses. Further, despite widespread effects on DNAm only seven studies accounted for smoking (Silva & Kamens, 2021). Smoking effects on DNAm are also dose- and time-dependent and anxious smokers commonly smoke more and for longer durations than non-anxious smokers (Fluharty et al., 2017; Philibert et al., 2014). We do note no CpGs consistently associated with smoking in the broader literature, nor the genes they annotate to (e.g., cg05575921 in *AHRR*, etc.), were significantly associated with anxiety in the MWASs reviewed here (Silva & Kamens, 2021) suggesting smoking was likely adequately accounted for.

Alternatively, limited cross-study replication may indicate some ANX-associated DNAm is not shared among ANX (i.e., DNAm is diagnosis-specific). For example, a MWAS of SAD and panic disorder found SAD, but not panic disorder, was associated with DNAm in *ANAPC16* (Ohi et al., 2024). This could indicate ANX-associated DNAm in *ANAPC16* is relevant only in SAD. A clearer difference in diagnosis-specific DNAm emerged when methylation risk scores (MRSs) were examined. MRSs are weighted sums of DNAm at associated CpGs that estimate methylomic risk for an outcome. The MRS examined in the panic disorder sample was generated from CpGs identified in the SAD MWAS, and vice versa (Ohi et al., 2024). While the MRS based on panic disorder was not significantly associated with SAD, the MRS based on SAD was greater in panic disorder cases. This suggests that while some DNAm related to SAD is potentially relevant to panic disorder, DNAm related to panic disorder may be less relevant to SAD. Some DNAm specific to each ANX could drive or result from their defining symptoms (e.g., spontaneous panic in panic disorder) and might reflect differences in underlying biological mechanisms across disorders. Not all ANX-associated DNAm is likely to be disorder-specific though, considering 3 MWASs of composite ANX phenotypes all identified methylome-wide significant associations (Domschke et al., 2025; Hettema et al., 2023; Kwon et al., 2025). Multivariate twin studies also report substantial overlap in genetic risk across ANX (Eley et al., 2008; Hettema et al., 2005), however such studies could not parse out the effects of epigenetic mechanisms on sources of variance. Thus, DNAm that overlaps multiple ANX could broadly contribute to ANX risk and promote, or be promoted by, ANX symptoms common to multiple ANX (e.g., excessive fear, avoidance etc.). Taken together, some ANX-associated DNAm may be shared among multiple ANX while other associations may be disorder-specific. Additional comparisons between different ANX should be conducted to further evaluate this theory.

Though our review did not identify ANX-associated DNAm significantly replicating across studies, some CpGs and genes were implicated in multiple studies and more than one ANX. Three CpGs (i.e., cg03019505, cg25372841, and cg25947600) positively associated with GAD (Guo et al., 2022) while also negatively associating with a composite phenotype of panic and or/agoraphobia, SAD, and multiple phobic disorders (Domschke et al., 2025). This supports their relevance in multiple anxiety disorders yet highlights potential differences between fear-based anxiety captured in the composite and distress-related anxiety (e.g., GAD) that should be investigated further. DNAm at a CpG in *RIOK3* nominally associated with lifetime anxiety case status (Hettema et al., 2023). Differential DNAm of *RIOK3* was also found in Guo et al.’s (2022) singular case group MWAS of GAD and OCD, which supports the likely involvement of *RIOK3* in multiple ANX. As OCD is not defined as an ANX by the DSM-5, this may further indicate altered DNAm in *RIOK3* is relevant for other mental disorders.

### Sex-Specificity of Immune and Inflammation Dysregulation

Notably, several identified genes are involved in immune or inflammatory processes suggesting DNAm may potentially dysregulate these processes in ANX. Examples include *RIOK3*, a known mediator of the innate immune response (Takashima et al., 2015), and *SLC43A2*, which is essential for the survival of regulatory T-cells (Saini et al., 2022). Genes implicated by other studies provide further support for the potential dysregulation of immune/inflammatory processes, including *HSPB6*, a known immunomodulant (Zininga et al., 2018), and *STK32B*, which encodes a serine/threonine kinase that regulates T-cell activation (Finlay & Cantrell, 2011).

However, other studies indicated not all ANX-associated DNAm, or its potential downstream impacts to immune and inflammatory processes, are shared among the sexes. For example, a MWAS of panic disorder found no significant CpGs in their full sample (Iurato et al., 2017). But, a sex-stratified analysis found cg07308824 in the promotor of *HECA*, while not significant in males, was significantly hypermethylated in females and replicated in females of an independent sample. This could indicate differential DNAm of *HECA* is related specifically to panic disorder in females, which is approximately twice that of males (McLean et al., 2011; Pigott, 2003). Additionally, given the predominant expression of *HECA* in lymphoid tissues and posited role in mediating inflammation (*Tissue Expression of HECA - summary - The human protein atlas*, 2024), this suggests some disruptions to immune and inflammatory processes in panic disorder are female-specific. In Iurato et al. (2017), functional analysis in a third female-only sample suggested this may be the case, as DNAm at cg07308824 associated with *HECA* mRNA expression at baseline and decreased expression after dexamethasone administration to treat panic-associated inflammation.

A second example concerns cg12701571 in the promoter of *ASB1*, which contains a SOCS domain known to regulate cytokine signaling. This site was differentially methylated between participants with severe and no or mild GAD in a sample of both sexes (Emeny et al., 2018). This suggests potential downstream impacts to cytokine production promoted by differential methylation of *ASB1* should be similarly altered in both sexes. However, when assessing the interaction between Interleukin 18 (IL18), a pro-inflammatory cytokine, and severe GAD predicted DNAm at cg12701571, a significant interaction was only found in females. Mean IL18 levels were also reduced in severely anxious females when compared to females with minimal or no GAD. Collectively, these studies provide preliminary evidence of a broad dysregulation of immune and inflammatory processes related to ANX-associated DNAm. Future studies should further explore the potential for sex-specific downstream impacts of this dysregulation.

### Considerations for Future Studies

#### Ethnic Diversity

While we found the ethnic diversity of samples used in MWASs of ANX is comparable to, or more diverse, than the MWAS literature for other mental disorders, most included studies primarily focused on White participants. DNAm can differ between ethnicities and ancestries due to genetics and unique cultural influences (Elliott et al., 2022; Galanter et al., 2017). Methylation quantitative-trait loci (meQTLs) are genetic variants that influence DNAm at certain locations and may be specific to ancestral subgroups, yet only 75% of DNAm variation across ethnicities is explained by genetic variation (Smith et al., 2014). Environmental and sociocultural experiences that vary across ethnicities, such as racial discrimination, dietary habits, or parenting styles, may also explain remaining differences(Chan et al., 2023; Meloni et al., 2022). ANX-associated CpGs and their effect sizes may differ between White and non-White individuals, bringing the generalizability of ANX-associated DNAm in mostly White participants into question. For example, 40 CpGs significantly associated with panic disorder in a Japanese sample (Shimada-Sugimoto et al., 2017), while no associations with panic disorder were found in a German sample (Ziegler et al., 2019). Similarly, a MWAS of GAD in Han Chinese and German individuals returned noticeably distinct results, though this may also have been influenced by study-specific definitions of GAD (Emeny et al., 2018; Guo et al., 2022). Future studies should make a concerted effort to include more ethnically diverse participants, which would allow the identification of associations relevant to all ethnicities. Such shared associations would enable a better assessment of the potential clinical relevance of ANX-associated DNAm, as they would not be specific to certain ethnic subgroups.

#### Tissue Choice

All studies examined DNAm in blood, which is easily accessible and thus constitutes an apt material when searching for biomarkers (Abi-Dargham et al., 2023); however, studies in brain tissue are needed to better our understanding of ANX-associated DNAm in the etiological tissue. In brain tissue, ANX-associated DNAm could be examined in both live and postmortem samples. However, the removal of live brain tissue solely for research purposes is highly unethical. Further, many regions of interest in ANX are inaccessible due to a high potential for surgery-related impacts on essential cognitive functions. Live brain samples can be obtained from participants undergoing medically necessary surgeries for severe brain-based disorders (e.g., epilepsy, cancer). These disorders, however, have their own widespread DNAm alterations. Postmortem samples could instead be obtained from a brain bank that stores and distributes brain tissue for research purposes. However, brain samples from individuals with mental disorders are rarely obtained by most brain banks, and, when they are, ANX are often not assessed. Low tissue availability would likely result in small sample sizes underpowered to detect relevant DNAm effects. Additionally, brain banks typically collect little to no information about disease onset, course and progression, which may restrict brain-based DNAm studies of ANX to only diagnostic status. Also, postmortem brain tissue degrades from death until preservation and may continue to degrade if stored in sub-optimal conditions (Ferrer et al., 2008). As such, their molecular signatures may not fully reflect processes in living tissues (Collado-Torres et al., 2023). Despite these challenges, studies in brain are needed to better our understanding of ANX-associated DNAm. Additionally, brain-based studies would allow evaluation of additional forms of methylation (i.e., hydroxymethylation and DNAm at non-CpG sites) that are prevalent in brain, but not other tissues, and may play vital roles in ANX.

Peripheral tissues, such as blood and saliva, have been suggested as proxy tissues for DNAm in brain. Support for this idea comes from average DNAm levels aggregated across all CpGs correlating at relatively high levels between live human brain tissue and blood, saliva, and/or buccal cells (*r* range: 0.84 to 0.91) (Braun et al., 2019; Nishitani et al., 2023; Zillich et al., 2022). However, these correlations weaken considerably when examined on an individual CpG level, with the strongest correlations seen in a limited number of CpGs exhibiting covarying DNAm (Hannon et al., 2015). A study of disease-related overlap between brain and other tissues in depression, a common comorbidity of ANX, produced similar results (Aberg et al., 2020). ANX-associated DNAm is thus expected to be different between brain and peripheral tissues, though there may be overlap at a small number of CpGs. No MWASs of ANX have been conducted in tissues other than blood, so we are unable to assess the suitability of using peripheral tissues as a proxy for brain in DNAm studies of ANX. To identify the overlap in potentially relevant tissues, MWASs in tissues other than blood need to be conducted and the concordance of ANX-associated DNAm between tissues assessed.

#### Cell-type Specificity

Regardless of the tissue used, DNAm at the same CpG in each cell-type of a tissue may vary significantly (Reinius et al., 2012). Yet only one study examined cell-type specific associations (Hettema et al., 2023). This study found whole blood may not capture all relevant ANX-associated DNAm. Only one CpG in *APC2* significantly associated with ANX in whole blood, while 280 and 82 CpG sites in CD14^+^ monocytes and CD15^+^ granulocytes, respectively, were differentially methylated. They also found differing ANX-associated DNAm between cell-types may have diverse downstream effects, as different processes were implicated in pathway analyses of different blood cell types. This study highlights the need for future research to further explore cell-type specific DNAm associations with ANX.

There are several available methods to assess cell-type specific DNAm. DNAm can be directly assayed in individual cells through single-cell DNAm sequencing. However, single-cell methods detect DNAm at only ∼5% of all potentially methylated CpGs and the specific CpGs assayed can vary significantly between cell-types (Cai et al., 2024), making single-cell DNAm sequencing challenging to compare across cell-types. Bulk tissues (e.g., brain, whole blood) could instead be sorted into selected cell-types (e.g., through FACS) and then assayed in each cell-type. However, if the number of samples is large, as is likely needed to detect the potentially small effect sizes of ANX-associated DNAm, the costs will grow exponentially with the number of cell-types selected. Also, not all samples and tissues can be sorted (e.g., blood spots) and a significant amount of material may be lost during sorting, even in tissues optimal for the technique (Cossarizza et al., 2017). Epigenomic deconvolution methods can be used to infer cell-type specific DNAm profiles from bulk tissue DNAm. This approach may be more applicable in large-scale studies as it can make use of extant data. Some deconvolution methods use reference panels of DNAm generated from pure, isolated cell-types to generate DNAm estimates (Salas et al., 2018; Teschendorff et al., 2017). However, these panels tend to include only major cell-types, limiting what can be learned about DNAm in less abundant cell-types, and may not be available for all tissues of potential interest. Reference-free methods are also available, though these approaches may produce less accurate cell-type specific estimates (Jeong et al., 2022). Any of the above methods would enable future studies to continue characterizing the role of cell-type specific DNAm in ANX, however the specific technique used should be chosen after careful consideration of its inherent limitations.

#### Longitudinal Study Designs

Lastly, we are unable to identify the temporal ordering or direction of effect of ANX-associated DNAm (i.e., does ANX precede changes in DNAm or vice versa?). Changes in DNAm can promote their own downstream effects, yet studies have also found DNAm changes can occur downstream of transcription factor binding or alterations in gene expression (Deplancke et al., 2016; Martin-Trujillo et al., 2020; Tseng et al., 2021). Knowing if ANX-associated DNAm contributes to disease development or is promoted by it heavily impacts its potential clinical relevance. Should DNAm changes precede ANX onset they may be useful predictive biomarkers or therapeutic targets in preventive or early intervention efforts. If ANX manifestation instead promotes changes in DNAm, ANX-associated DNAm would have more utility in diagnostics and monitoring disease course. Carefully designed longitudinal studies establishing if DNAm drives ANX, or results from it, must be conducted. Standardized collection protocols should be used to obtain multiple phenotypic and DNAm measurements over time, with measurement beginning before ANX onset. Ideally, enough measurements should be taken to evaluate both short- and long-term effects of ANX and DNAm. Additionally, collection of phenotypic information should be as comprehensive as possible and include known confounders of DNAm.

#### Phenotype Harmonization

The included studies used a variety of ANX phenotypes from recent symptoms of generalized anxiety (Ciuculete et al., 2018) to individual ANX to composite ANX status. While twin and genetic association studies have demonstrated substantial overlap in genetic risk factors across ANX (Eley et al., 2008; Hettema et al., 2005; Meier & Deckert, 2019), the degree of overlap in their DNAm is unknown. This represents a possible cryptic source of heterogeneity between studies using different phenotypes. DNAm associations with lifetime- and current-ANX status may also differ, yet one MWAS found high correlations between analyses conducted on participants with lifetime ANX versus those with current ANX (Hettema et al., 2023). The optimum design of future studies needs to consider closer alignment of study phenotypes.

### Limitations of the Current Review

First, we limited our review to studies examining associations with DNAm assessed on a methylome-wide scale. Thus, we excluded candidate gene studies with their limited focus on DNAm in promoter regions and a high likelihood of false positives. However, we note none of the commonly studied candidate genes (e.g., *SLC6A4*, *FKBP5*, *OXTR*, *NR3C1*) were among significant findings in MWAS of ANX (Table S1). We also did not examine the relationship between ANX and other epigenetic modifications (e.g., histone modifications, non-coding RNAs), for which few to no studies in humans exist despite evidence from animal studies supporting their involvement in ANX- and stress-related phenotypes (Ell et al., 2024). Third, due to substantial variations in study design, PRISMA guidelines suggested the currently available literature is not sufficiently homogeneous to conduct a meta-analysis. Our ability to standardize information from individual studies to create a more uniform dataset for meta-analysis was further restricted by limited access to the raw data itself. As such, we are unable to further quantify the strength of evidence regarding ANX-associated DNAm. Regardless, our review was thorough, provides meaningful insight into key themes and limitations of the literature, and offers clear suggestions to enhance our understanding of ANX-associated DNAm.

## Conclusion

The literature reviewed here lays the groundwork for future discoveries by providing initial evidence of ANX-associated DNAm in humans. Research examining the relationship between DNAm and ANX continues to expand and evolve, buoyed by advances in technology and a growing acknowledgment of the widespread and detrimental impacts of ANX. We are optimistic more replicable findings will be identified as sample sizes increase and limitations we have noted are addressed.

## Data Availability

All data produced in the present work are contained in the manuscript

## References

1. Aberg, K. A., Dean, B., Shabalin, A. A., Chan, R. F., Han, L. K. M., Zhao, M., van Grootheest, G., Xie, L. Y., Milaneschi, Y., Clark, S. L., Turecki, G., Penninx, B., & van den Oord, E. (2020). Methylome-wide association findings for major depressive disorder overlap in blood and brain and replicate in independent brain samples. Mol Psychiatry, 25(6), 1344–1354. 10.1038/s41380-018-0247-6

2. Abi-Dargham, A., Moeller, S. J., Ali, F., DeLorenzo, C., Domschke, K., Horga, G., Jutla, A., Kotov, R., Paulus, M. P., Rubio, J. M., Sanacora, G., Veenstra-VanderWeele, J., & Krystal, J. H. (2023). Candidate biomarkers in psychiatric disorders: state of the field. World Psychiatry, 22(2), 236–262. 10.1002/wps.21078

3. Alisch, R. S., Van Hulle, C., Chopra, P., Bhattacharyya, A., Zhang, S. C., Davidson, R. J., Kalin, N. H., & Goldsmith, H. H. (2017). A multi-dimensional characterization of anxiety in monozygotic twin pairs reveals susceptibility loci in humans. Transl Psychiatry, 7(12), 1282. 10.1038/s41398-017-0047-9

4. American Psychiatric Association. (2013). Diagnostic and statistical manual of mental disorders (5th ed.). 10.1176/appi.books.9780890425596

5. Border, R., Johnson, E. C., Evans, L. M., Smolen, A., Berley, N., Sullivan, P. F., & Keller, M. C. (2019). No Support for Historical Candidate Gene or Candidate Gene-by-Interaction Hypotheses for Major Depression Across Multiple Large Samples. Am J Psychiatry, 176(5), 376–387. 10.1176/appi.ajp.2018.18070881

6. Bortoluzzi, A., Salum, G. A., da Rosa, E. D., Chagas, V. S., Castro, M. A. A., & Manfro, G. G. (2018). DNA methylation in adolescents with anxiety disorder: a longitudinal study. Sci Rep, 8(1), 13800. 10.1038/s41598-018-32090-1

7. Braun, P. R., Han, S., Hing, B., Nagahama, Y., Gaul, L. N., Heinzman, J. T., Grossbach, A. J., Close, L., Dlouhy, B. J., Howard, M. A., 3rd, Kawasaki, H., Potash, J. B., & Shinozaki, G. (2019). Genome-wide DNA methylation comparison between live human brain and peripheral tissues within individuals. Transl Psychiatry, 9(1), 47. 10.1038/s41398-019-0376-y

8. Cai, M., Zhou, J., McKennan, C., & Wang, J. (2024). scMD facilitates cell type deconvolution using single-cell DNA methylation references. Commun Biol, 7(1), 1. 10.1038/s42003-023-05690-5

9. Campagna, M. P., Xavier, A., Lechner-Scott, J., Maltby, V., Scott, R. J., Butzkueven, H., Jokubaitis, V. G., & Lea, R. A. (2021). Epigenome-wide association studies: current knowledge, strategies and recommendations. Clin Epigenetics, 13(1), 214. 10.1186/s13148-021-01200-8

10. Chagnon, Y. C., Potvin, O., Hudon, C., & Preville, M. (2015). DNA methylation and single nucleotide variants in the brain-derived neurotrophic factor (BDNF) and oxytocin receptor (OXTR) genes are associated with anxiety/depression in older women. Front Genet, 6, 230. 10.3389/fgene.2015.00230

11. Chan, M. H., Merrill, S. M., Konwar, C., & Kobor, M. S. (2023). An integrative framework and recommendations for the study of DNA methylation in the context of race and ethnicity. Discov Soc Sci Health, 3(1), 9. 10.1007/s44155-023-00039-z

12. Ciuculete, D. M., Bostrom, A. E., Tuunainen, A. K., Sohrabi, F., Kular, L., Jagodic, M., Voisin, S., Mwinyi, J., & Schioth, H. B. (2018). Changes in methylation within the STK32B promoter are associated with an increased risk for generalized anxiety disorder in adolescents. J Psychiatr Res, 102, 44–51. 10.1016/j.jpsychires.2018.03.008

13. Collado-Torres, L., Klei, L., Liu, C., Kleinman, J. E., Hyde, T. M., Geschwind, D. H., Gandal, M. J., Devlin, B., & Weinberger, D. R. (2023). Comparison of gene expression in living and postmortem human brain. medRxiv. 10.1101/2023.11.08.23298172

14. Cossarizza, A., Chang, H. D., Radbruch, A., Akdis, M., Andra, I., Annunziato, F., Bacher, P., Barnaba, V., Battistini, L., Bauer, W. M., Baumgart, S., Becher, B., Beisker, W., Berek, C., Blanco, A., Borsellino, G., Boulais, P. E., Brinkman, R. R., Buscher, M., … Zimmermann, J. (2017). Guidelines for the use of flow cytometry and cell sorting in immunological studies. Eur J Immunol, 47(10), 1584–1797. 10.1002/eji.201646632

15. Craske, M. G., Rauch, S. L., Ursano, R., Prenoveau, J., Pine, D. S., & Zinbarg, R. E. (2009). What is an anxiety disorder? Depress Anxiety, 26(12), 1066–1085. 10.1002/da.20633

16. Danielsdottir, H. B., Aspelund, T., Shen, Q., Halldorsdottir, T., Jakobsdottir, J., Song, H., Lu, D., Kuja-Halkola, R., Larsson, H., Fall, K., Magnusson, P. K. E., Fang, F., Bergstedt, J., & Valdimarsdottir, U. A. (2024). Adverse Childhood Experiences and Adult Mental Health Outcomes. JAMA Psychiatry, 81(6), 586–594. 10.1001/jamapsychiatry.2024.0039

17. Deplancke, B., Alpern, D., & Gardeux, V. (2016). The Genetics of Transcription Factor DNA Binding Variation. Cell, 166(3), 538–554. 10.1016/j.cell.2016.07.012

18. Doering, S., Lichtenstein, P., Gillberg, C., Ntr, Middeldorp, C. M., Bartels, M., Kuja-Halkola, R., & Lundstrom, S. (2019). Anxiety at age 15 predicts psychiatric diagnoses and suicidal ideation in late adolescence and young adulthood: results from two longitudinal studies. BMC Psychiatry, 19(1), 363. 10.1186/s12888-019-2349-3

19. Domschke, K., Schiele, M. A., Crespo Salvador, O., Zillich, L., Lipovsek, J., Pittig, A., Heinig, I., Ridderbusch, I. C., Straube, B., Richter, J., Hollandt, M., Plag, J., Fydrich, T., Koelkebeck, K., Weber, H., Lueken, U., Dannlowski, U., Margraf, J., Schneider, S., … Deckert, J. (2025). Epigenetic markers of disease risk and psychotherapy response in anxiety disorders - a longitudinal analysis of the DNA methylome. Mol Psychiatry. 10.1038/s41380-025-03038-5

20. Domschke, K., Tidow, N., Kuithan, H., Schwarte, K., Klauke, B., Ambree, O., Reif, A., Schmidt, H., Arolt, V., Kersting, A., Zwanzger, P., & Deckert, J. (2012). Monoamine oxidase A gene DNA hypomethylation - a risk factor for panic disorder? Int J Neuropsychopharmacol, 15(9), 1217–1228. 10.1017/S146114571200020X

21. Drzewiecki, C. M., & Fox, A. S. (2024). Understanding the heterogeneity of anxiety using a translational neuroscience approach. Cogn Affect Behav Neurosci, 24(2), 228–245. 10.3758/s13415-024-01162-3

22. Ehrlich, M., & Lacey, M. (2013). DNA methylation and differentiation: silencing, upregulation and modulation of gene expression. Epigenomics, 5(5), 553–568. 10.2217/epi.13.43

23. Eley, T. C., Rijsdijk, F. V., Perrin, S., O’Connor, T. G., & Bolton, D. (2008). A multivariate genetic analysis of specific phobia, separation anxiety and social phobia in early childhood. J Abnorm Child Psychol, 36(6), 839–848. 10.1007/s10802-008-9216-x

24. Ell, M. A., Schiele, M. A., Iovino, N., & Domschke, K. (2024). Epigenetics of Fear, Anxiety and Stress - Focus on Histone Modifications. Curr Neuropharmacol, 22(5), 843–865. 10.2174/1570159X21666230322154158

25. Elliott, H. R., Burrows, K., Min, J. L., Tillin, T., Mason, D., Wright, J., Santorelli, G., Davey Smith, G., Lawlor, D. A., Hughes, A. D., Chaturvedi, N., & Relton, C. L. (2022). Characterisation of ethnic differences in DNA methylation between UK-resident South Asians and Europeans. Clin Epigenetics, 14(1), 130. 10.1186/s13148-022-01351-2

26. Emeny, R. T., Baumert, J., Zannas, A. S., Kunze, S., Wahl, S., Iurato, S., Arloth, J., Erhardt, A., Balsevich, G., Schmidt, M. V., Weber, P., Kretschmer, A., Pfeiffer, L., Kruse, J., Strauch, K., Roden, M., Herder, C., Koenig, W., Gieger, C., … Ladwig, K. H. (2018). Anxiety Associated Increased CpG Methylation in the Promoter of Asb1: A Translational Approach Evidenced by Epidemiological and Clinical Studies and a Murine Model. Neuropsychopharmacology, 43(2), 342–353. 10.1038/npp.2017.102

27. Farlik, M., Halbritter, F., Muller, F., Choudry, F. A., Ebert, P., Klughammer, J., Farrow, S., Santoro, A., Ciaurro, V., Mathur, A., Uppal, R., Stunnenberg, H. G., Ouwehand, W. H., Laurenti, E., Lengauer, T., Frontini, M., & Bock, C. (2016). DNA Methylation Dynamics of Human Hematopoietic Stem Cell Differentiation. Cell Stem Cell, 19(6), 808–822. 10.1016/j.stem.2016.10.019

28. Ferrer, I., Martinez, A., Boluda, S., Parchi, P., & Barrachina, M. (2008). Brain banks: benefits, limitations and cautions concerning the use of post-mortem brain tissue for molecular studies. Cell Tissue Bank, 9(3), 181–194. 10.1007/s10561-008-9077-0

29. Finlay, D., & Cantrell, D. (2011). The coordination of T-cell function by serine/threonine kinases. Cold Spring Harb Perspect Biol, 3(1), a002261. 10.1101/cshperspect.a002261

30. Fluharty, M., Taylor, A. E., Grabski, M., & Munafo, M. R. (2017). The Association of Cigarette Smoking With Depression and Anxiety: A Systematic Review. Nicotine Tob Res, 19(1), 3–13. 10.1093/ntr/ntw140

31. Galanter, J. M., Gignoux, C. R., Oh, S. S., Torgerson, D., Pino-Yanes, M., Thakur, N., Eng, C., Hu, D., Huntsman, S., Farber, H. J., Avila, P. C., Brigino-Buenaventura, E., LeNoir, M. A., Meade, K., Serebrisky, D., Rodriguez-Cintron, W., Kumar, R., Rodriguez-Santana, J. R., Seibold, M. A., … Zaitlen, N. (2017). Differential methylation between ethnic sub-groups reflects the effect of genetic ancestry and environmental exposures. Elife, 6. 10.7554/eLife.20532

32. Gottschalk, M. G., & Domschke, K. (2017). Genetics of generalized anxiety disorder and related traits. Dialogues Clin Neurosci, 19(2), 159–168. 10.31887/DCNS.2017.19.2/kdomschke

33. Guo, L., Ni, Z., Wei, G., Cheng, W., Huang, X., & Yue, W. (2022). Epigenome-wide DNA methylation analysis of whole blood cells derived from patients with GAD and OCD in the Chinese Han population. Transl Psychiatry, 12(1), 465. 10.1038/s41398-022-02236-x

34. Hannon, E., Lunnon, K., Schalkwyk, L., & Mill, J. (2015). Interindividual methylomic variation across blood, cortex, and cerebellum: implications for epigenetic studies of neurological and neuropsychiatric phenotypes. Epigenetics, 10(11), 1024–1032. 10.1080/15592294.2015.1100786

35. Hettema, J. M., Prescott, C. A., Myers, J. M., Neale, M. C., & Kendler, K. S. (2005). The structure of genetic and environmental risk factors for anxiety disorders in men and women. Arch Gen Psychiatry, 62(2), 182–189. 10.1001/archpsyc.62.2.182

36. Hettema, J. M., van den Oord, E., Zhao, M., Xie, L. Y., Copeland, W. E., Penninx, B., Aberg, K. A., & Clark, S. L. (2023). Methylome-wide association study of anxiety disorders. Mol Psychiatry, 28(8), 3484–3492. 10.1038/s41380-023-02205-w

37. Houseman, E. A., Kim, S., Kelsey, K. T., & Wiencke, J. K. (2015). DNA Methylation in Whole Blood: Uses and Challenges. Curr Environ Health Rep, 2(2), 145–154. 10.1007/s40572-015-0050-3

38. Howe, A. S., Buttenschon, H. N., Bani-Fatemi, A., Maron, E., Otowa, T., Erhardt, A., Binder, E. B., Gregersen, N. O., Mors, O., Woldbye, D. P., Domschke, K., Reif, A., Shlik, J., Koks, S., Kawamura, Y., Miyashita, A., Kuwano, R., Tokunaga, K., Tanii, H., … De Luca, V. (2016). Candidate genes in panic disorder: meta-analyses of 23 common variants in major anxiogenic pathways. Mol Psychiatry, 21(5), 665–679. 10.1038/mp.2015.138

39. Iurato, S., Carrillo-Roa, T., Arloth, J., Czamara, D., Diener-Holzl, L., Lange, J., Muller-Myhsok, B., Binder, E. B., & Erhardt, A. (2017). “DNA Methylation signatures in panic disorder”. Transl Psychiatry, 7(12), 1287. 10.1038/s41398-017-0026-1

40. Javaid, S. F., Hashim, I. J., Hashim, M. J., Stip, E., Samad, M. A., & Ahbabi, A. A. (2023). Epidemiology of anxiety disorders: global burden and sociodemographic associations. Middle East Current Psychiatry, 30(1), 44. 10.1186/s43045-023-00315-3

41. Jeong, Y., de Andrade, E. S. L. B., Thalmeier, D., Toth, R., Ganslmeier, M., Breuer, K., Plass, C., & Lutsik, P. (2022). Systematic evaluation of cell-type deconvolution pipelines for sequencing-based bulk DNA methylomes. Brief Bioinform, 23(4). 10.1093/bib/bbac248

42. Johnson, E. C., Border, R., Melroy-Greif, W. E., de Leeuw, C. A., Ehringer, M. A., & Keller, M. C. (2017). No Evidence That Schizophrenia Candidate Genes Are More Associated With Schizophrenia Than Noncandidate Genes. Biol Psychiatry, 82(10), 702–708. 10.1016/j.biopsych.2017.06.033

43. Kalin, N. H. (2020). The Critical Relationship Between Anxiety and Depression. Am J Psychiatry, 177(5), 365–367. 10.1176/appi.ajp.2020.20030305

44. Kessler, R. C., Chiu, W. T., Demler, O., Merikangas, K. R., & Walters, E. E. (2005). Prevalence, severity, and comorbidity of 12-month DSM-IV disorders in the National Comorbidity Survey Replication. Arch Gen Psychiatry, 62(6), 617–627. 10.1001/archpsyc.62.6.617

45. Kwon, Y., Blazyte, A., Jeon, Y., Kim, Y. J., An, K., Jeon, S., Ryu, H., Shin, D. H., Ahn, J., Um, H., Kang, Y., Bak, H., Kim, B. C., Lee, S., Jung, H. T., Shin, E. S., & Bhak, J. (2025). Identification of 17 novel epigenetic biomarkers associated with anxiety disorders using differential methylation analysis followed by machine learning-based validation. Clin Epigenetics, 17(1), 24. 10.1186/s13148-025-01819-x

46. Levey, D. F., Gelernter, J., Polimanti, R., Zhou, H., Cheng, Z., Aslan, M., Quaden, R., Concato, J., Radhakrishnan, K., Bryois, J., Sullivan, P. F., Million Veteran, P., & Stein, M. B. (2020). Reproducible Genetic Risk Loci for Anxiety: Results From approximately 200,000 Participants in the Million Veteran Program. Am J Psychiatry, 177(3), 223–232. 10.1176/appi.ajp.2019.19030256

47. Li, M., D’Arcy, C., Li, X., Zhang, T., Joober, R., & Meng, X. (2019). What do DNA methylation studies tell us about depression? A systematic review. Transl Psychiatry, 9(1), 68. 10.1038/s41398-019-0412-y

48. Loyfer, N., Magenheim, J., Peretz, A., Cann, G., Bredno, J., Klochendler, A., Fox-Fisher, I., Shabi-Porat, S., Hecht, M., Pelet, T., Moss, J., Drawshy, Z., Amini, H., Moradi, P., Nagaraju, S., Bauman, D., Shveiky, D., Porat, S., Dior, U., … Kaplan, T. (2023). A DNA methylation atlas of normal human cell types. Nature, 613(7943), 355–364. 10.1038/s41586-022-05580-6

49. Mansell, G., Gorrie-Stone, T. J., Bao, Y., Kumari, M., Schalkwyk, L. S., Mill, J., & Hannon, E. (2019). Guidance for DNA methylation studies: statistical insights from the Illumina EPIC array. BMC Genomics, 20(1), 366. 10.1186/s12864-019-5761-7

50. Martin-Trujillo, A., Patel, N., Richter, F., Jadhav, B., Garg, P., Morton, S. U., McKean, D. M., DePalma, S. R., Goldmuntz, E., Gruber, D., Kim, R., Newburger, J. W., Porter, G. A., Jr., Giardini, A., Bernstein, D., Tristani-Firouzi, M., Seidman, J. G., Seidman, C. E., Chung, W. K., … Sharp, A. J. (2020). Rare genetic variation at transcription factor binding sites modulates local DNA methylation profiles. PLoS Genet, 16(11), e1009189. 10.1371/journal.pgen.1009189

51. McLean, C. P., Asnaani, A., Litz, B. T., & Hofmann, S. G. (2011). Gender differences in anxiety disorders: prevalence, course of illness, comorbidity and burden of illness. J Psychiatr Res, 45(8), 1027–1035. 10.1016/j.jpsychires.2011.03.006

52. Meier, S. M., & Deckert, J. (2019). Genetics of Anxiety Disorders. Curr Psychiatry Rep, 21(3), 16. 10.1007/s11920-019-1002-7

53. Meloni, M., Moll, T., Issaka, A., & Kuzawa, C. W. (2022). A biosocial return to race? A cautionary view for the postgenomic era. Am J Hum Biol, 34(7), e23742. 10.1002/ajhb.23742

54. Morrison, F. G., Miller, M. W., Logue, M. W., Assef, M., & Wolf, E. J. (2019). DNA methylation correlates of PTSD: Recent findings and technical challenges. Prog Neuropsychopharmacol Biol Psychiatry, 90, 223–234. 10.1016/j.pnpbp.2018.11.011

55. Nelson, E. C., Heath, A. C., Madden, P. A., Cooper, M. L., Dinwiddie, S. H., Bucholz, K. K., Glowinski, A., McLaughlin, T., Dunne, M. P., Statham, D. J., & Martin, N. G. (2002). Association between self-reported childhood sexual abuse and adverse psychosocial outcomes: results from a twin study. Arch Gen Psychiatry, 59(2), 139–145. 10.1001/archpsyc.59.2.139

56. Nishitani, S., Isozaki, M., Yao, A., Higashino, Y., Yamauchi, T., Kidoguchi, M., Kawajiri, S., Tsunetoshi, K., Neish, H., Imoto, H., Arishima, H., Kodera, T., Fujisawa, T. X., Nomura, S., Kikuta, K., Shinozaki, G., & Tomoda, A. (2023). Cross-tissue correlations of genome-wide DNA methylation in Japanese live human brain and blood, saliva, and buccal epithelial tissues. Transl Psychiatry, 13(1), 72. 10.1038/s41398-023-02370-0

57. Ohi, K., Fujikane, D., Takai, K., Kuramitsu, A., Muto, Y., Sugiyama, S., & Shioiri, T. (2024). Epigenetic signatures of social anxiety, panic disorders and stress experiences: Insights from genome-wide DNA methylation risk scores. Psychiatry Res, 337, 115984. 10.1016/j.psychres.2024.115984

58. Page, M. J., McKenzie, J. E., Bossuyt, P. M., Boutron, I., Hoffmann, T. C., Mulrow, C. D., Shamseer, L., Tetzlaff, J. M., Akl, E. A., Brennan, S. E., Chou, R., Glanville, J., Grimshaw, J. M., Hrobjartsson, A., Lalu, M. M., Li, T., Loder, E. W., Mayo-Wilson, E., McDonald, S., … Moher, D. (2021). The PRISMA 2020 statement: An updated guideline for reporting systematic reviews. Int J Surg, 88, 105906. 10.1016/j.ijsu.2021.105906

59. Petersen, C. L., Chen, J. Q., Salas, L. A., & Christensen, B. C. (2020). Altered immune phenotype and DNA methylation in panic disorder. Clin Epigenetics, 12(1), 177. 10.1186/s13148-020-00972-9

60. Philibert, R. A., Beach, S. R., & Brody, G. H. (2014). The DNA methylation signature of smoking: an archetype for the identification of biomarkers for behavioral illness. Nebr Symp Motiv, 61, 109–127. 10.1007/978-1-4939-0653-6_6

61. Pigott, T. A. (2003). Anxiety disorders in women. Psychiatr Clin North Am, 26(3), 621–672, vi-vii. 10.1016/s0193-953x(03)00040-6

62. Purves, K. L., Coleman, J. R. I., Meier, S. M., Rayner, C., Davis, K. A. S., Cheesman, R., Baekvad-Hansen, M., Borglum, A. D., Wan Cho, S., Jurgen Deckert, J., Gaspar, H. A., Bybjerg-Grauholm, J., Hettema, J. M., Hotopf, M., Hougaard, D., Hubel, C., Kan, C., McIntosh, A. M., Mors, O., … Eley, T. C. (2020). A major role for common genetic variation in anxiety disorders. Mol Psychiatry, 25(12), 3292–3303. 10.1038/s41380-019-0559-1

63. Qi, L., & Teschendorff, A. E. (2022). Cell-type heterogeneity: Why we should adjust for it in epigenome and biomarker studies. Clin Epigenetics, 14(1), 31. 10.1186/s13148-022-01253-3

64. Reinius, L. E., Acevedo, N., Joerink, M., Pershagen, G., Dahlen, S. E., Greco, D., Soderhall, C., Scheynius, A., & Kere, J. (2012). Differential DNA methylation in purified human blood cells: implications for cell lineage and studies on disease susceptibility. PLoS One, 7(7), e41361. 10.1371/journal.pone.0041361

65. Saini, N., Naaz, A., Metur, S. P., Gahlot, P., Walvekar, A., Dutta, A., Davathamizhan, U., Sarin, A., & Laxman, S. (2022). Methionine uptake via the SLC43A2 transporter is essential for regulatory T-cell survival. Life Sci Alliance, 5(12). 10.26508/lsa.202201663

66. Salas, L. A., Koestler, D. C., Butler, R. A., Hansen, H. M., Wiencke, J. K., Kelsey, K. T., & Christensen, B. C. (2018). An optimized library for reference-based deconvolution of whole-blood biospecimens assayed using the Illumina HumanMethylationEPIC BeadArray. Genome Biol, 19(1), 64. 10.1186/s13059-018-1448-7

67. Schiele, M. A., Gottschalk, M. G., & Domschke, K. (2020). The applied implications of epigenetics in anxiety, affective and stress-related disorders - A review and synthesis on psychosocial stress, psychotherapy and prevention. Clin Psychol Rev, 77, 101830. 10.1016/j.cpr.2020.101830

68. Shabalin, A. A., Aberg, K. A., & van den Oord, E. J. (2015). Candidate gene methylation studies are at high risk of erroneous conclusions. Epigenomics, 7(1), 13–15. 10.2217/epi.14.70

69. Shimada-Sugimoto, M., Otowa, T., Miyagawa, T., Umekage, T., Kawamura, Y., Bundo, M., Iwamoto, K., Tochigi, M., Kasai, K., Kaiya, H., Tanii, H., Okazaki, Y., Tokunaga, K., & Sasaki, T. (2017). Epigenome-wide association study of DNA methylation in panic disorder. Clin Epigenetics, 9, 6. 10.1186/s13148-016-0307-1

70. Silva, C. P., & Kamens, H. M. (2021). Cigarette smoke-induced alterations in blood: A review of research on DNA methylation and gene expression. Exp Clin Psychopharmacol, 29(1), 116–135. 10.1037/pha0000382

71. Smith, A. K., Kilaru, V., Kocak, M., Almli, L. M., Mercer, K. B., Ressler, K. J., Tylavsky, F. A., & Conneely, K. N. (2014). Methylation quantitative trait loci (meQTLs) are consistently detected across ancestry, developmental stage, and tissue type. BMC Genomics, 15, 145. 10.1186/1471-2164-15-145

72. Sullivan, P. F. (2007). Spurious genetic associations. Biol Psychiatry, 61(10), 1121–1126. 10.1016/j.biopsych.2006.11.010

73. Takashima, K., Oshiumi, H., Takaki, H., Matsumoto, M., & Seya, T. (2015). RIOK3-mediated phosphorylation of MDA5 interferes with its assembly and attenuates the innate immune response. Cell Rep, 11(2), 192–200. 10.1016/j.celrep.2015.03.027

74. Teschendorff, A. E., Breeze, C. E., Zheng, S. C., & Beck, S. (2017). A comparison of reference-based algorithms for correcting cell-type heterogeneity in Epigenome-Wide Association Studies. BMC Bioinformatics, 18(1), 105. 10.1186/s12859-017-1511-5

75. Tissue Expression of HECA - summary - The human protein atlas. (2024). Human Protein Atlas. Retrieved August 16 from https://www.proteinatlas.org/ENSG00000112406-HECA/tissue

76. Tseng, C. C., Wong, M. C., Liao, W. T., Chen, C. J., Lee, S. C., Yen, J. H., & Chang, S. J. (2021). Genetic Variants in Transcription Factor Binding Sites in Humans: Triggered by Natural Selection and Triggers of Diseases. Int J Mol Sci, 22(8). 10.3390/ijms22084187

77. Wiegand, A., Kreifelts, B., Munk, M. H. J., Geiselhart, N., Ramadori, K. E., MacIsaac, J. L., Fallgatter, A. J., Kobor, M. S., & Nieratschker, V. (2021). DNA methylation differences associated with social anxiety disorder and early life adversity. Transl Psychiatry, 11(1), 104. 10.1038/s41398-021-01225-w

78. Womersley, J. S., Roeh, S., Martin, L., Ahmed-Leitao, F., Sauer, S., Rex-Haffner, M., Hemmings, S. M. J., Binder, E. B., & Seedat, S. (2022). FKBP5 intron 7 methylation is associated with higher anxiety proneness and smaller right thalamus volume in adolescents. Brain Struct Funct, 227(8), 2809–2820. 10.1007/s00429-022-02577-9

79. Zhou, Q., Jackson-Cook, C., Lyon, D., Perera, R., & Archer, K. J. (2015). Identifying molecular features associated with psychoneurological symptoms in women with breast cancer using multivariate mixed models. Cancer Inform, 14(Suppl 2), 139–145. 10.4137/CIN.S17276

80. Ziegler, C., Dannlowski, U., Brauer, D., Stevens, S., Laeger, I., Wittmann, H., Kugel, H., Dobel, C., Hurlemann, R., Reif, A., Lesch, K. P., Heindel, W., Kirschbaum, C., Arolt, V., Gerlach, A. L., Hoyer, J., Deckert, J., Zwanzger, P., & Domschke, K. (2015). Oxytocin receptor gene methylation: converging multilevel evidence for a role in social anxiety. Neuropsychopharmacology, 40(6), 1528–1538. 10.1038/npp.2015.2

81. Ziegler, C., Grundner-Culemann, F., Schiele, M. A., Schlosser, P., Kollert, L., Mahr, M., Gajewska, A., Lesch, K. P., Deckert, J., Kottgen, A., & Domschke, K. (2019). The DNA methylome in panic disorder: a case-control and longitudinal psychotherapy-epigenetic study. Transl Psychiatry, 9(1), 314. 10.1038/s41398-019-0648-6

82. Zillich, L., Poisel, E., Streit, F., Frank, J., Fries, G. R., Foo, J. C., Friske, M. M., Sirignano, L., Hansson, A. C., Nothen, M. M., Witt, S. H., Walss-Bass, C., Spanagel, R., & Rietschel, M. (2022). Epigenetic Signatures of Smoking in Five Brain Regions. J Pers Med, 12(4). 10.3390/jpm12040566

83. Zininga, T., Ramatsui, L., & Shonhai, A. (2018). Heat Shock Proteins as Immunomodulants. Molecules, 23(11). 10.3390/molecules23112846

84. Zou, Z., Zhang, Y., Huang, Y., Wang, J., Min, W., Xiang, M., Zhou, B., & Li, T. (2023). Integrated genome-wide methylation and expression analyses provide predictors of diagnosis and early response to antidepressant in panic disorder. J Affect Disord, 322, 146–155. 10.1016/j.jad.2022.10.049

